# Spread of COVID-19 through Georgia, USA. Near-term projections and impacts of social distancing via a metapopulation model

**DOI:** 10.1101/2020.05.28.20115642

**Authors:** Stephen J. Beckett, Marian Dominguez-Mirazo, Seolha Lee, Clio Andris, Joshua S. Weitz

**Affiliations:** School of Biological Sciences, Georgia Institute of Technology, Atlanta, GA, USA; Center for Microbial Dynamics and Infection, Georgia Institute of Technology, Atlanta, GA, USA; School of City & Regional Planning, Georgia Institute of Technology, Atlanta, GA, USA; School of Interactive Computing, Georgia Institute of Technology, Atlanta, GA, USA; School of Physics, Georgia Institute of Technology, Atlanta, GA, USA

## Abstract

Epidemiological forecasts of COVID-19 spread at the country and/or state level have helped shape public health interventions. However, such models leave a scale-gap between the spatial resolution of actionable information (i.e. the county or city level) and that of modeled viral spread. States and nations are not spatially homogeneous and different areas may vary in disease risk and severity. For example, COVID-19 has age-stratified risk. Similarly, ICU units, PPE and other vital equipment are not equally distributed within states. Here, we implement a county-level epidemiological framework to assess and forecast COVID-19 spread through Georgia, where 1,933 people have died from COVID-19 and 44,638 cases have been documented as of May 27, 2020. We find that county-level forecasts trained on heterogeneity due to clustered events can continue to predict epidemic spread over multiweek periods, potentially serving efforts to prepare medical resources, manage supply chains, and develop targeted public health interventions. We find that the premature removal of physical (social) distancing could lead to rapid increases in cases or the emergence of sustained plateaus of elevated fatalities.

**Highlights:**

- Contact heterogeneity and age-structured risk is represented in a metapopulation model.
- The metapopulation model can be used in the near-term to make joint predictions on cases, hospitalizations, and fatalities at county levels.
- In Georgia, a long-term plateauing epidemic, rather than a distinctive epidemic peak, is possible.

## 1 Introduction

COVID-19 is a global pandemic which has (as of May 27, 2020) caused more than five and a half million reported cases and over 350,000 reported fatalities worldwide. Empirical evidence suggests that a large proportion of cases are asymptomatic [1, 2] making transmission an often ‘invisible’ occurrence which increases challenges for disease control efforts. Public health responses to help curb transmission of COVID-19 have far-reaching health, economic, social and emotional consequences, and depend on epidemiological models. However, these models are often developed at large spatial scales that do not necessarily incorporate local information about factors such as population demographics, travel patterns, or ICU capacity [3, 4]. We seek to model this heterogeneity to better understand how COVID-19 will be transmitted across spatial scales that can be translated into local surveillance and action. Our effort is similar in spirit to multiple models that also aim to bring actionable information to local scales (e.g., [5, 6, 7, 8, 9]).

To tailor models to locales, we extended an age-structured epidemiological model with eight epidemiological states, including sub-acute and critical hospitalizations (as in [10, 11]). The use of an age-specific model is essential given that the elderly are more severely impacted [12, 10, 13]; additional comorbidities include obesity and environmental factors which could be incorporated into future work. We extend this model to incorporate space and spatial population structure [14] using a metapopulation modeling framework that implicitly accounts for transport between county-level ‘patches’, by assuming that the mixing population can be broken into two categories: the local patch population; and a commuter patch population. We term this the Metapopulation A Ge-structured Epidemiological (MAGE) model. While this framework could be applied generically, in this work we focus on modeling the spread of COVID-19 through counties in Georgia, where previous work predicts that thousands of deaths could occur [11]. However, such state-level models do not capture local commuting patterns, or availability of resources and crisis-support that will be useful to inform public health interventions.

As of May 27, 2020 over 1,933 people have died from COVID-19 in the state of Georgia, with over 44,638 recorded cases, and these numbers will soon be surpassed and outdated. The first recorded case was in Metropolitan Atlanta in early March, but the city of Albany in Southern Georgia also became a hotspot of cases after a large extended family and their friends contracted the disease [15]. COVID-19 has spread across the whole state (Figure 1, absolute (top) and per capita (bottom) cumulative cases). While Atlanta and Albany are hotspots for COVID-19, Southwestern Georgia as a whole is disproportionately affected when normalized by population size. This heterogeneity in infection rates indicates a need to understand community transmission at local scales – this is particularly relevant in Georgia which has 159 counties and a population of 10.6 million residents. It is also important to note that recorded cases are only a proxy for COVID-19 spread as there is a delay between taking a test and the results being recorded. Additionally, many of those in Georgia who have been exposed to COVID-19 have not been tested – because they were ineligible, were unable due to disparities in testing effort [16], or unwilling to seek tests; or were asymptomatic and did not realize they were carrying the disease. As a result, the number of actual cases is almost certainly much higher than records show. Here, we use reported cases and fatalities, county demographics and county commuter patterns, coupled with a mechanistic mathematical model, to project COVID-19 spread at the county-level in Georgia and to evaluate the benefits of interventions, focusing on sustained efforts to reduce infection via different levels of physical (social) distancing.

**Figure 1:**
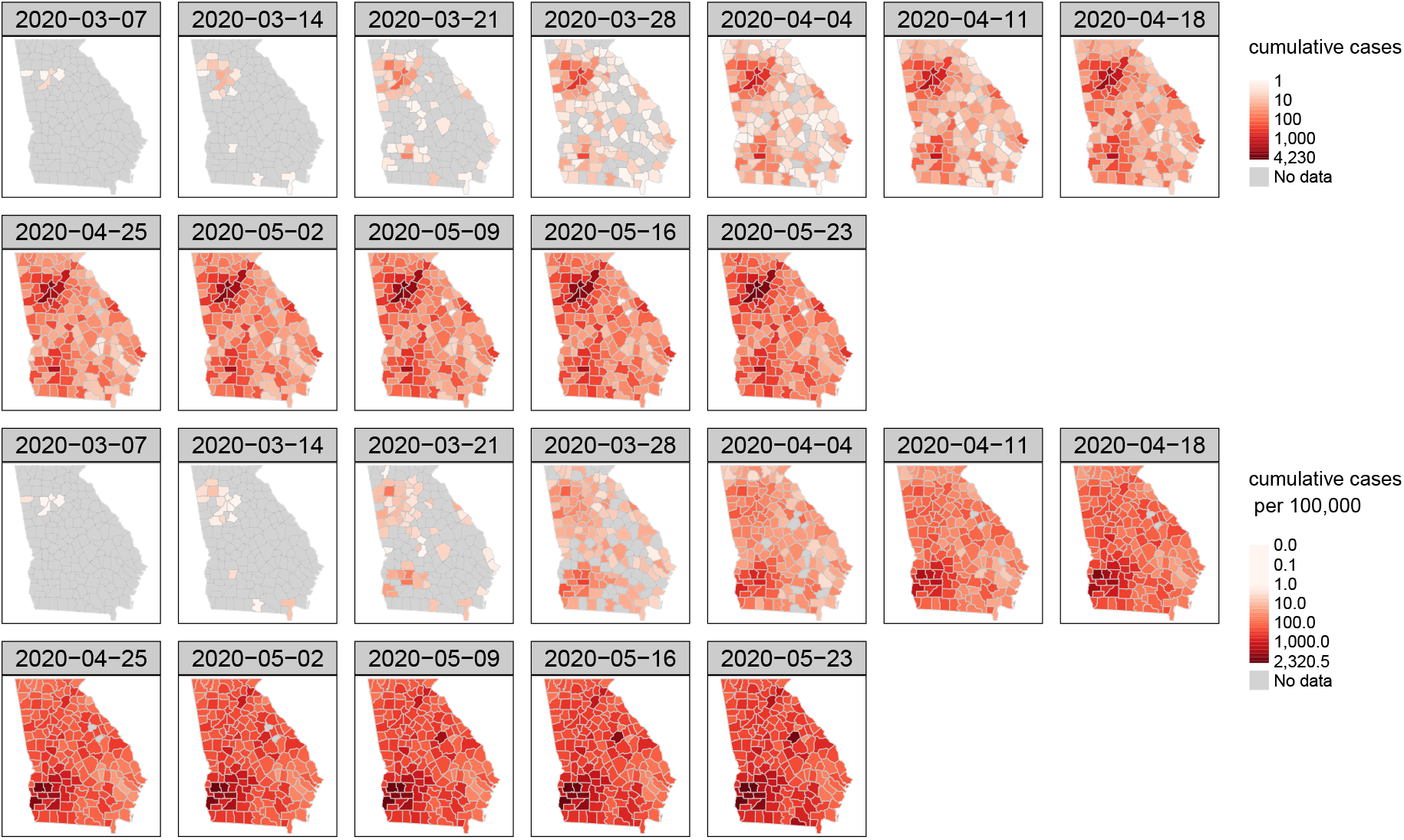
Weekly time-lapse of cumulative recorded cases of COVID-19 data. Top: cumulative recorded case in each county. Bottom: cumulative case data per 100,000 people in Georgia counties. Uses data obtained from The New York Times county-level COVID-19 dataset, based on reports from state and local health agencies.

## 2 Methods

### 2.1 Mathematical model of COVID-19 spatial spread

We adapt existing epidemiological compartment models [10, 12] to model the spatial spread of COVID-19. We model COVID-19 spread through Georgia using 159 connected patches (i.e. counties), where individuals are classified as being in one of eight epidemiological compartments. Five of the eight epidemiological states are allowed to move, and potentially transmit the disease between counties: susceptible *S*, exposed *E*, infected asymptomatic *I_A_*, infected symptomatic *I_S_*, and recovered *R*. Additionally, we consider that some symptomatic cases require hospital care that can be sub-acute *I_h,sub_* or critical *I_h,crit_*, where critical hospitalization may result in death *D*. Furthermore, individuals are stratified by age – we consider nine age classes: 0–9, 10–19, 20–29, 30–39, 40–49, 50–59, 60–69, 70–79, and over 80. This model has a total of 159 *×* 8 *×* 9 = 11, 488 variables. To model the spatial spread of COVID-19 we assume that local individuals spend their time mixing with either the local population; or with a commuting population, which accounts for commuting between counties. We define the commuter patch population as the local population who stay within a patch, minus those who leave to visit other patches, plus those who travel into the patch from other patches. The relative time spent mixing with either “local” or “commuting” populations is mediated by the weights *w_L_* and *w_C_*. Commutes are used to signify movement between counties, although much of the population is no longer commuting during lockdowns. To account for this we mediate transmission rates by a physical distancing (a.k.a. social distancing) factor *κ*. The model is visualized in Figure 2. The system of nonlinear differential equations governing this metapopulation model with age-structured epidemiology can be described for patch *i* and age *a* as follows:

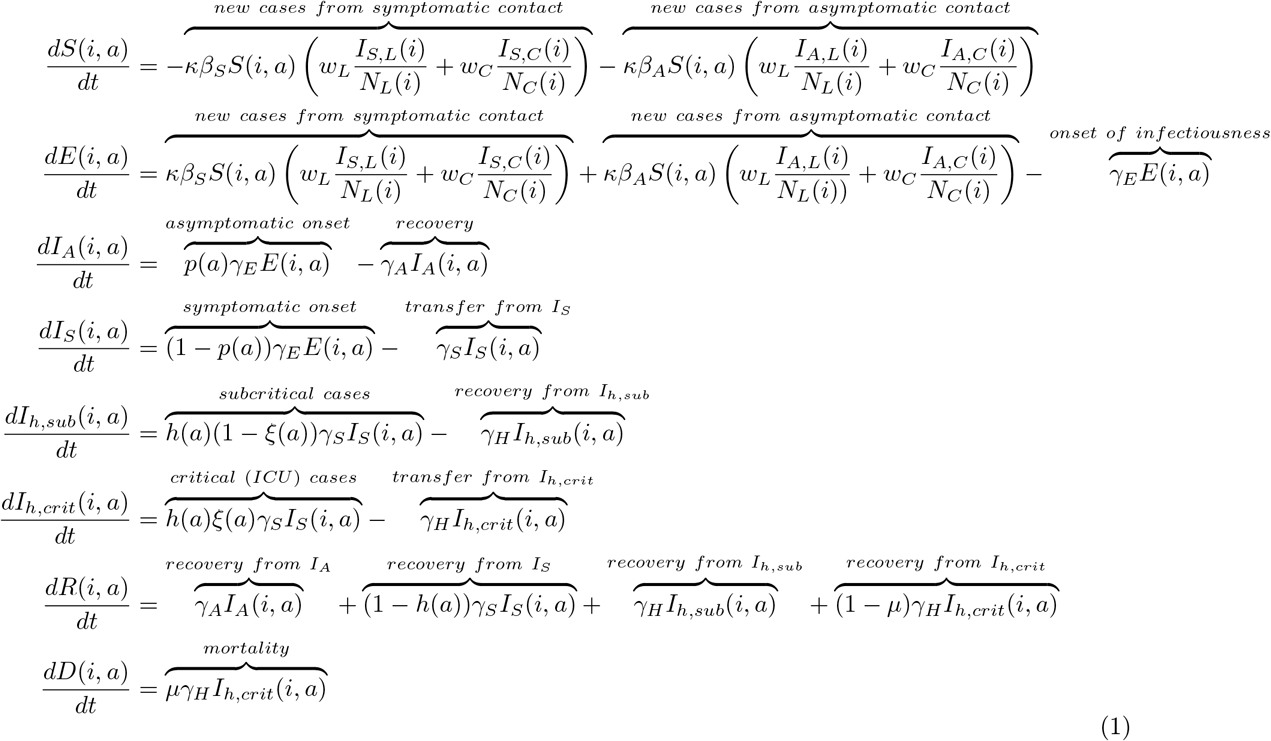

**Figure 2:**
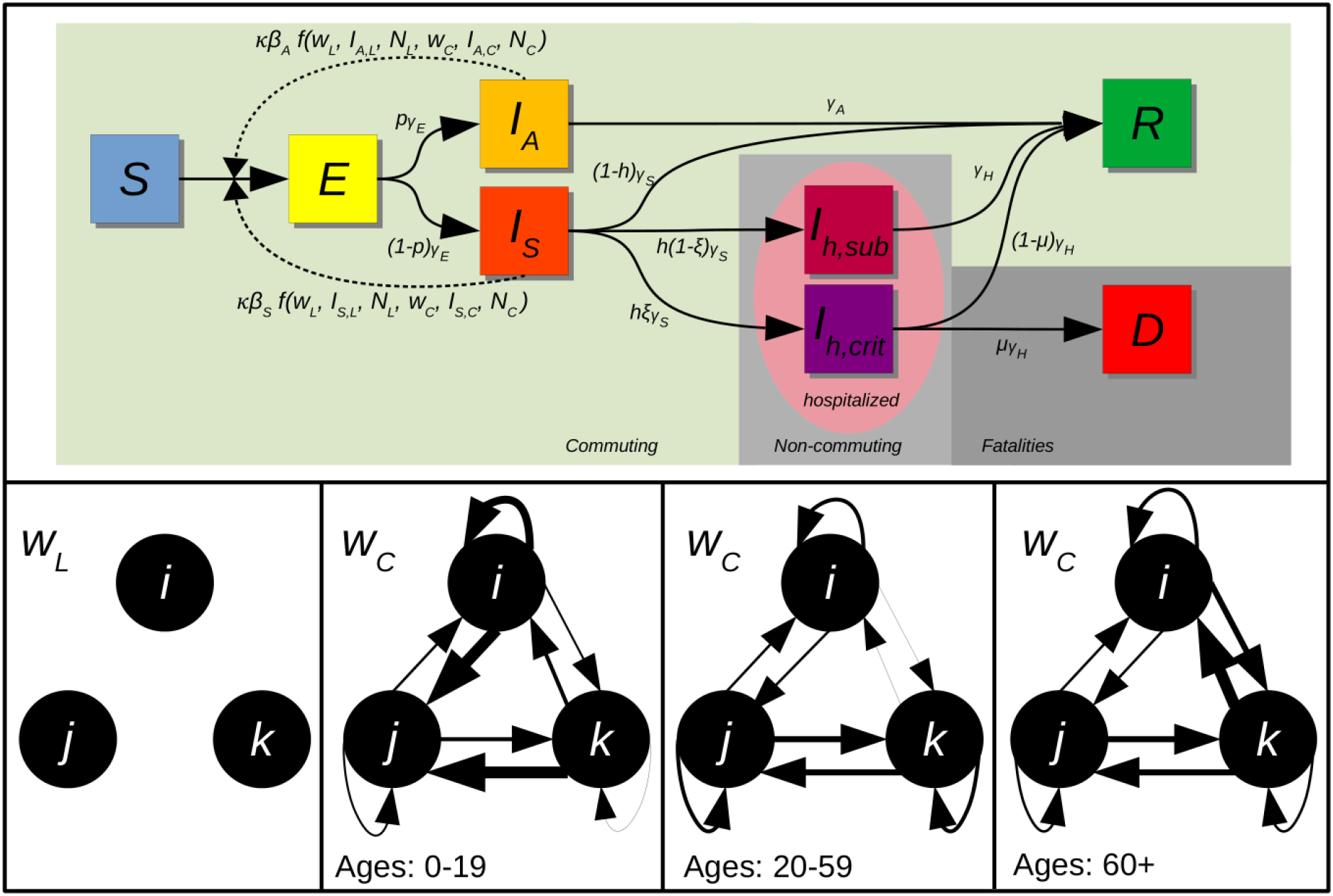
Schematic of the compartmental MAGE model with commuting. Top: The epidemic model and transition pathways within a single patch and for a single age group. Susceptible (*S*) individuals interact with local asymptomatic (*I_A,L_*) and symptomatic (*I_S,L_*) infected individuals from the local population (*N_L_*) *w_L_* of the time; or with commuting asymptomatic (*I_A,C_*) and symptomatic (*I_S,C_*) infected individuals from the commuting population (*N_C_*) *w_C_* of the time and become exposed (*E*). Exposed individuals either experience the onset of the disease asymptomatically (*I_A_*) or symptomatically(*I_S_*). A subgroup of symptomatically infected individuals will require hospital care with either sub-acute (*I_h,sub_*) or critical (*I_h,crit_*) severity. A subset of critical cases will die (*D*). Those who recover (*R*) are assumed to develop immunity on outbreak timescales i.e. at least a year. Hospitalized cases and those who are dead are not part of the commuting population that can move between patches. Bottom: subpanels show commuting patterns for the *S,E,I_A_,I_S_* and *R* states between patches *i,j* and *k*. For a relative time *w_L_* individuals interact only with the local population, e.g., when at or close to home. For the relative time *w_C_* individuals are mixing within a commuter population which is potentially heterogeneous with respect to age (here by three age classes). The commuter population calculates net migration to the patch from the local population.

where *I_S,L_* and *I_S,C_* are the total number (across all age classes) of symptomatic infectious individuals; *I_A,L_* and *I_A,C_* are the total number of asymptomatic infectious individuals; and *N_L_* and *N_C_* are the total number of living individuals during “local” (L) and “commuting” (C) times respectively. During the relative time spent in the local phase:

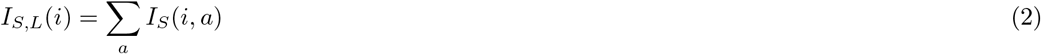

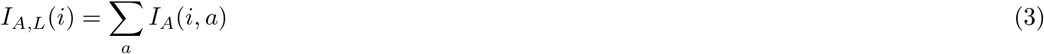

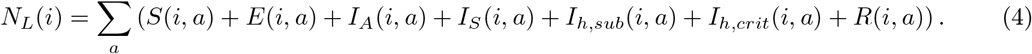

To capture commutes we introduce the transport matrix *M* that contains the proportion of people who move from patch *i* to patch *j*. We describe how the transport matrix is constructed in Supporting Information S2. Diagonal elements of *M* are negative and describe the proportion of people who leave the current patch. The current patch is indexed by the column elements; such that each column sums to 0. Hospitalized or deceased individuals are removed from the commuting population. Thus, the commuter populations for each patch can be computed by multiplying the transport matrix (159×159) by a vector of a particular age and epidemiological state across all nodes (159×1) to find the net population change during commuter times (159×1). We denote these net changes with hats (an example calculation is shown in the Supporting Information equation S6). We can then calculate the commuting population components as:

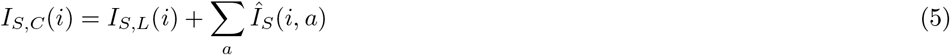

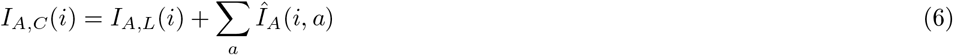

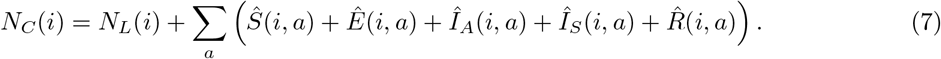

Parameters are chosen based on the Imperial College London Model [10] (Tables 1 and 2).

**Table 1:** Epidemiological characteristics.

| Parameter | Meaning | Value |
| --- | --- | --- |
| $\beta_a$ | Asymptomatic transmission rate | 0.3/day |
| $\beta_s$ | Symptomatic transmission rate | 0.6/day |
| $1/\gamma_E$ | Mean exposed period | 4 days |
| $1/\gamma_A$ | Mean asymptomatic period | 6 days |
| $1/\gamma_S$ | Mean symptomatic period | 6 days |
| $1/\gamma_H$ | Mean hospital period | 10 days |
| $w_L$ | Relative time in local phase | 2/3 |
| $w_C$ | Relative time in commuting phase | 1/3 |
| $\kappa$ | Relative reduction in transmission rates | var. |

**Table 2:** Age-stratified risk for COVID-19. Of note, the model assumes that 50% of ICU cases die. Parameters in line with [12, 10].

| Age class | Frac. Asymptomatic ($p$) | Hospital Frac. ($h$) | ICU (given hospitalization) Frac. ($\xi$) |
| --- | --- | --- | --- |
| 0-9 | 0.95 | 0.001 | 0.05 |
| 10-19 | 0.95 | 0.003 | 0.05 |
| 20-29 | 0.9 | 0.012 | 0.05 |
| 30-39 | 0.8 | 0.032 | 0.05 |
| 40-49 | 0.7 | 0.049 | 0.063 |
| 50-59 | 0.6 | 0.102 | 0.122 |
| 60-69 | 0.4 | 0.166 | 0.274 |
| 70-79 | 0.2 | 0.243 | 0.432 |
| 80+ | 0.2 | 0.273 | 0.709 |

### 2.2 Epidemiological simulations

In order to make county-level projections, we use data released by the Georgia Department of Public Health on the March 28, 2020, around the time social distancing measures were introduced, to set county-level initial conditions in the model. We use these initial conditions to run the model in an evaluation phase (March 28 – April 28). The model is recalibrated and projected forward from April 28 in an assessment phase. Given the ongoing need to respond rapidly to COVID-19, we released our assessments on April 28th as an unpublished report [17]. All simulations here take these same parameters as the basis for further analysis.

#### 2.2.1 Evaluation Phase

We initialize an outbreak with a single person aged 20–29 in a Georgia-wide model (spatial features removed (see equation S1), and simulate the epidemiological dynamics forward in time with no reduction in transmission from baseline levels (*κ* = 1) until the number of recorded deaths at the state level on March 28, 2020 is reached. We then assign this distribution to the county level proportional to the number of deaths recorded in each county. Counties with no recorded deaths are assigned an equally distributed portion of deaths whose reporting county is unknown. We then simulate the MAGE model forward in time under a scenario of 50% transmission reduction due to physical (social) distancing interventions (*κ* = 0.5) through April 28, 2020.

#### 2.2.2 Assessment Phase

We recalibrate the model on April 28, 2020 against Georgia Department of Public Health data for April 28, 2020. We do this by simulating epidemiological dynamics with the Georgia-wide model in the same manner as in the evaluation phase to March 28, 2020. We then run the Georgia-wide model forward from March 28 to April 28 with a 50% reduction in transmission rates (*κ* = 0.5) until the number of recorded deaths at the state level on April 28, 2020 is reached. As before, we assign this distribution to the county level proportional to the number of deaths recorded in each county. Counties without recorded deaths are assigned an equally distributed portion of deaths whose reporting county is unknown. We forecast the MAGE model forward from April 28 to May 1 under a scenario of 50% transmission reduction to May 1. Following May 1, we project three scenarios forward in time assuming social distancing policies result in 0%, 50% or 75% reductions in transmission rates. As the timescale to initialize the model differs from the actual timescale, simulated cumulative numbers overestimate the number of cases that occur. We therefore adjust cumulative numbers of hospitalizations and cases to match recorded levels on April 28 for our projections.

### 2.3 Data and software availability

Georgia age-stratified population county data was obtained from the US Census Bureau American Community Survey for 2018 [18]. Commuting data was obtained from US Census LODES [19]. County-level COVID-19 case and death counts were obtained from the Georgia Department of Public Health COVID-19 7pm Daily Status Report (https://dph.georgia.gov/covid-19-daily-status-report); and from The New York Times county-level COVID-19 dataset, based on reports from state and local health agencies (https://github.com/nytimes/covid-19-data). We interpret deaths per day and cases per day as the increase in cumulative recorded totals between days. ICU capacity data was obtained from the Kaiser Health news analysis of hospital cost reports filed to the Centers for Medicare and Medicaid services. Aggregated regions in the excess capacity analysis are the regions of the Georgia Regional Coordinating Hospital (RCH) System (https://southhealthdistrict.com/programsservices/emergency-prep/healthcare-preparedness-coalitions/ The MAGE model was developed in Julia [20] and numerical integration was performed using a 5(4) order adaptive time-stepping method [21] implemented in the DifferentialEquations.jl [22] package. Visualizations were made using the R packages tmap [23] and sf [24]. All code for simulations and plotting is available at http://www.github.com/WeitzGroup/MAGEmodel_covid19_GA and is archived [25].

## 3 Results

### 3.1 Model-data fitting suggests significant ascertainment bias

Models were initialized using data for March 28, 2020. Simulations were projected forward to July 4. In our initial evaluation phase of the MAGE model (Figure 3), we find initial conditions that are similar to data with respect to fatality and cumulative hospitalizations, but differ in their description of cumulative exposed individuals. The MAGE model suggests that around 50,000 people had encountered COVID-19 by March 28, 2020. Down-projecting case numbers in Figure 3c suggests that there may be between 5 and 10 unrecorded cases for every recorded case. While these may appear to be large numbers we note these estimates are compatible with previous work on ‘NowCasting’ and ascertainment bias [26] and that these numbers are small relative to the total population in Georgia (<2% of the population). In Figure S1 we show alternative scenarios during the evaluation phase that show that the data seem closest to a scenario with a 50% reduction in transmission rates. This scenario most closely matches the observed steady increase in cumulative deaths and hospitalizations during the period between March 28 and April 28. Related data suggests that Georgia has reduced mobility by around 40–60% on average since March 28 [27, 28]. In the short term we find that physical (social) distancing may have already prevented thousands of deaths, as well as reduced hospital demand, and new case and death rates relative to baseline scenarios with no intervention (Figure S1).

**Figure 3:**
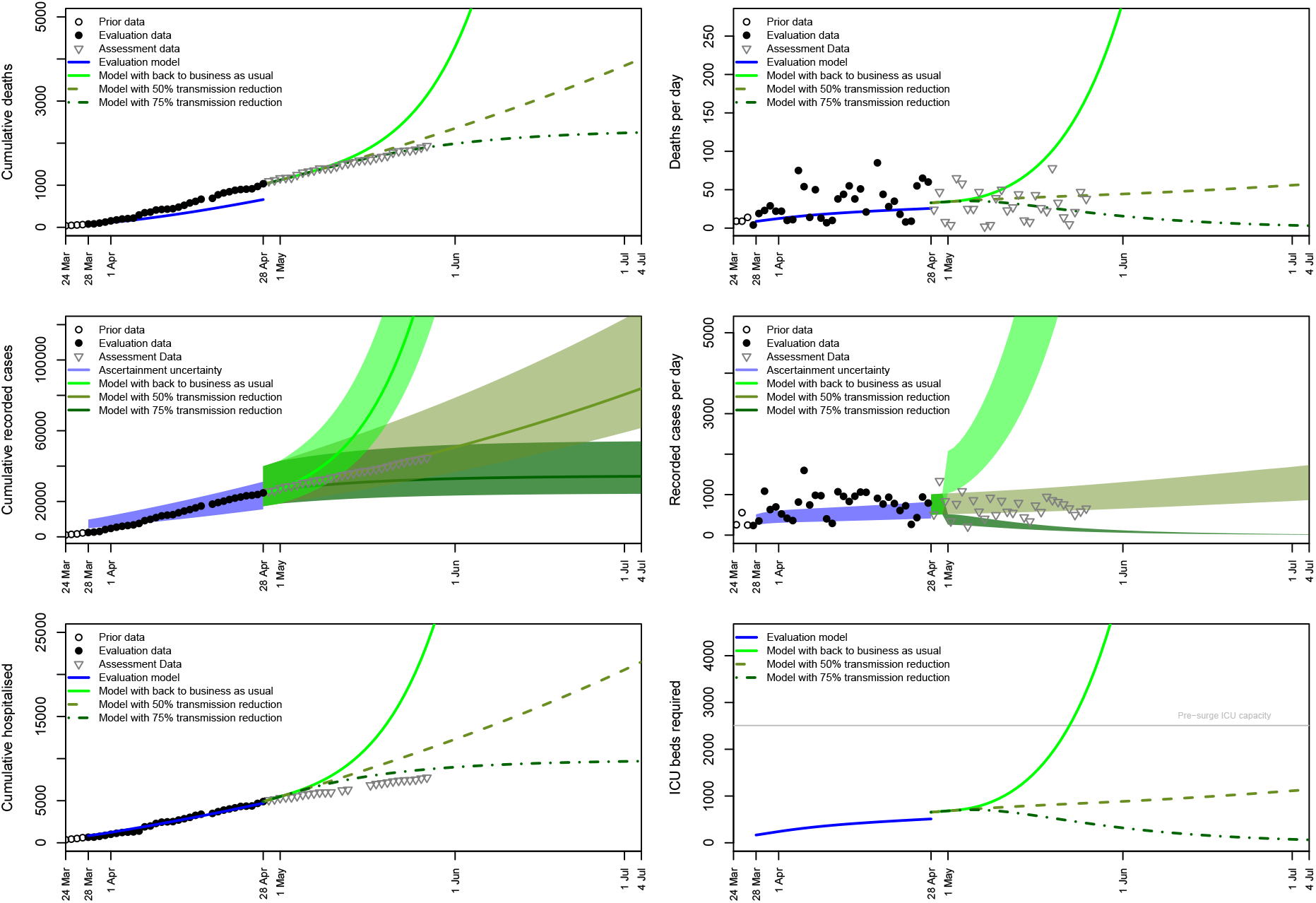
Short-term COVID-19 projections for the state of Georgia from March 28, 2020 to July 4, 2020. The evaluation model (with 50% social distancing) was initialized on March 28 and recalibrated on April 28 for projections to July 4. Forecasting scenarios represent a return to business as usual, continued intervention, or increased intervention. Case numbers are down-projected by a factor of 5–10*×* to compare with recorded data from testing. a) Cumulative deaths b) deaths per day c) cumulative cases recorded d) cases recorded per day e) cumulative hospitalizations f) number of ICU beds required. Pre-surge ICU bed capacity is highlighted with a horizontal line.

### 3.2 Short-term projections of COVID-19 cases and fatalities in Georgia

The MAGE model appears to underestimate fatalities in the evaluation phase. For this reason we recalibrated the model on April 28 and projected forward to July 4 under three scenarios for altered transmission dynamics. In projecting forward in the assessment phase, we estimate that returning to business as usual would lead to an exponential rise in cases, hospitalizations and fatalities. On the other hand, an increased intervention strategy, e.g., with 75% reduction in transmission rates could lead to a decline in cases, hospitalizations and deaths. While cumulative hospitalization data appears more consistent with a 75% transmission reduction scenario, recorded case data appears most consistent with a scenario with a 50% reduction in transmission rates in which cases, hospitalizations and fatalities gradually increase through time, consistent with little to no change in the effective social distancing parameter before vs. after the reopening.

### 3.3 Heterogeneous impacts on hospital responses

In Figure 4 we report cumulative deaths and projected cumulative deaths at the county level. Most fatalities occurred in Fulton County and Dougherty County, consistent with the locations with the highest reported case prevalence (see Figure 1). In contrast to recorded cases, many counties appear to have no COVID-19 related fatalities, as of May 27 2020. The distribution of cumulative deaths appears generally consistent with simulation projections, which contain assumed commuting structure. Similar spatial distributions are expected across the different assessment scenarios, but with varying intensity.

**Figure 4:**
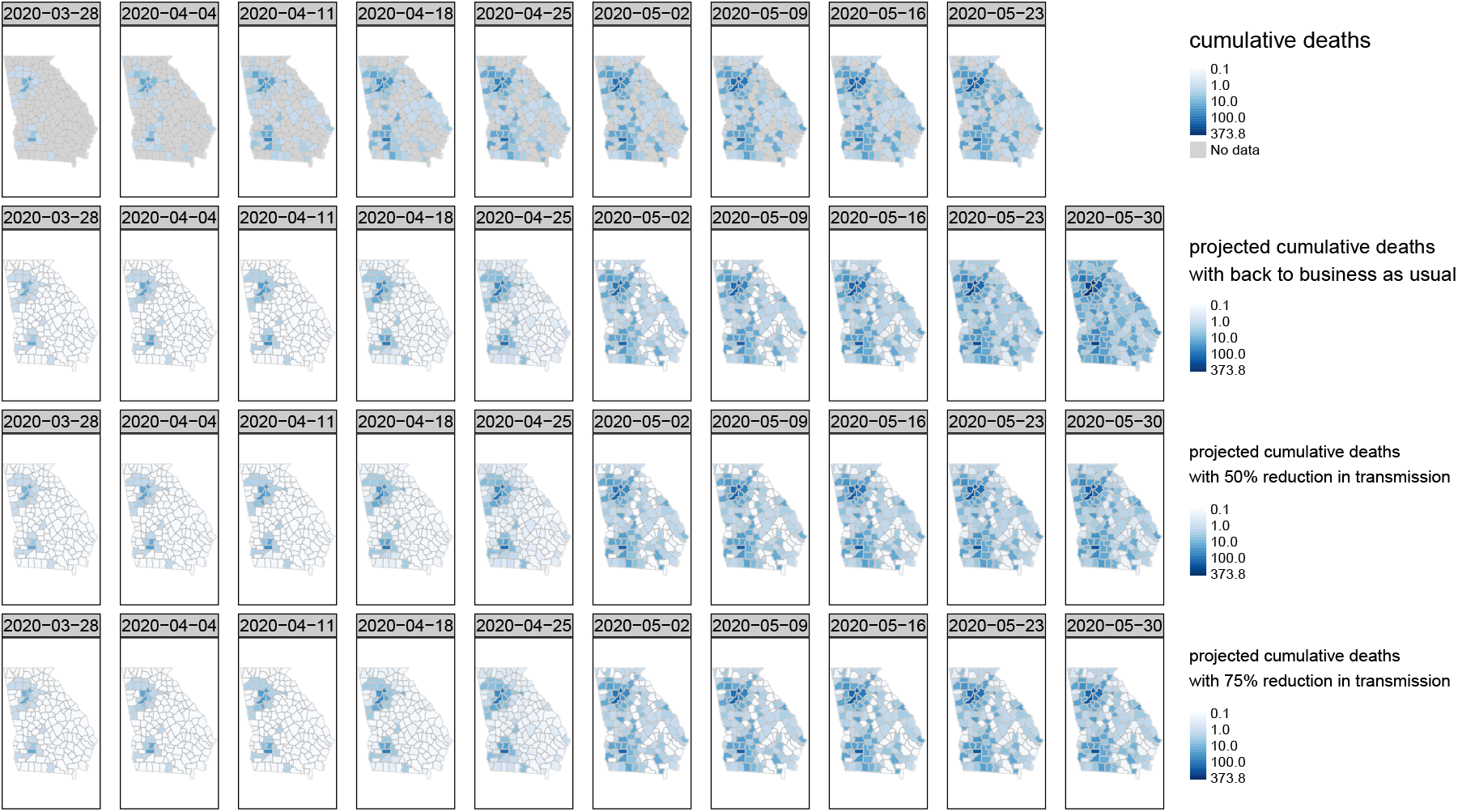
Weekly COVID-19 deaths in Georgia from March 28, 2020 to May 30, 2020. Top: cumulative recorded COVID-19 deaths. The bottom three rows show deaths from the evaluation model between March 28 and April 28. Projections from April 28; 2nd row: with a return to business as usual. 3rd row: with continued intervention as a 50% reduction in transmission rates. 4th row: with increased intervention as a 75% reduction in transmission rates.

To account for disparities in ICU availability we find the ratio of ICU demand to ICU capacity which is projected in Figure 5. As many counties do not have ICU units, we base our analysis of capacity relative to demand on Georgia’s Regional Coordinating Hospital System. In Figure 5, reports of values above one show that demand exceeds capacity in that region. We note that this analysis does not consider the use of ICU beds for non-COVID-19 cases, meaning that in reality ICU services are likely under even more pressure. Additionally, we do not consider the ability of hospitals to staff these beds. With a back to business as usual scenario with no physical (social) distancing interventions we find that almost all 14 regions would be above baseline capacity by the end of May. Demand would be nearly 5x pre-surge capacity in the Fairview Park Hospital (Central Georgia) and Phoebe Putney Memorial Hospital (Southwestern Georgia) regions and over 2x pre-surge capacity in the Hamilton Medical Center (Northwestern Georgia) and Wellstar Kennestone Hospital (Atlanta Metro West) regions. Our model also does not include dynamic feedback between available capacity and demand. This means that our baseline simulations may underestimate the severity of the impact of COVID-19 given additional fatalities that may occur when medical facilities are overstretched. In scenarios where social distancing reduces transmission by 50% we find that hospital systems would be more resilient within this timeframe with only the Phoebe Putney Memorial Hospital region (housing Albany, Georgia) exceeding capacity. While cases across Georgia are increasing in this scenario, we find that the Phoebe Putney Memorial Hospital region will surpass capacity first, followed by the Fairview Park Hospital region. We find that the Augusta University Medical Center region may be most prepared to meet demand within its region; and could be the focus of load-bearing efforts.

**Figure 5:**
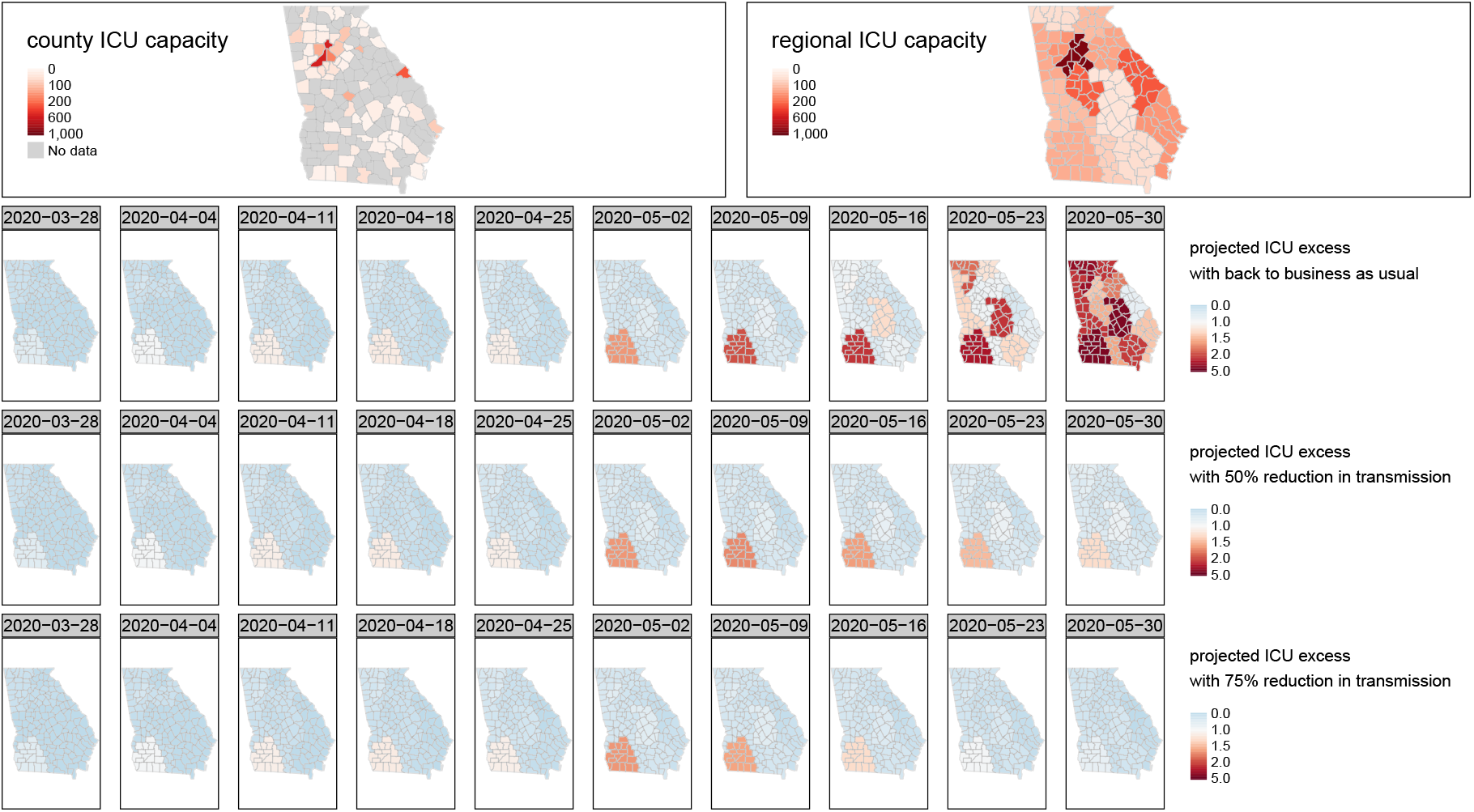
Projected excess ICU bed demand aggregated to hospital regions from March 28, 2020 to May 30, 2020. A value equal to one is the point at which demand meets capacity. A value of two means demand is double capacity. Top row shows ICU capacity across counties (left) and hospital regions (right). The bottom three rows show excess ICU bed demand from the evaluation model between March 28 and April 28. Projections from April 28; 2nd row: with a return to business as usual. 3rd row: with continued intervention as a 50% reduction in transmission rates. 4th row: with increased intervention as a 75% reduction in transmission rates.

### 3.4 Medium-term projections and impacts of social distancing scenarios

In the medium term it is useful to consider when things may return to business as usual [29]. In order to do this, we extend our projections to September 24, 2020 (Figure 6). If transmission rates were to return to pre-intervention levels in a back to business as usual scenario the epidemic may peak in mid-June – leading to mass fatalities. With extreme social distancing (more than current levels), new cases could decrease and the epidemic could curtail by late June, i.e., cases would be low enough such that input of cases from outside the state rather than community transmission would be the predominant factor in driving dynamics. Or, under a 50% reduction in transmission via social distancing the epidemic may plateau – where the numbers of inpatients vs. outpatients appears near balanced through and beyond September [30, 31].

**Figure 6:**
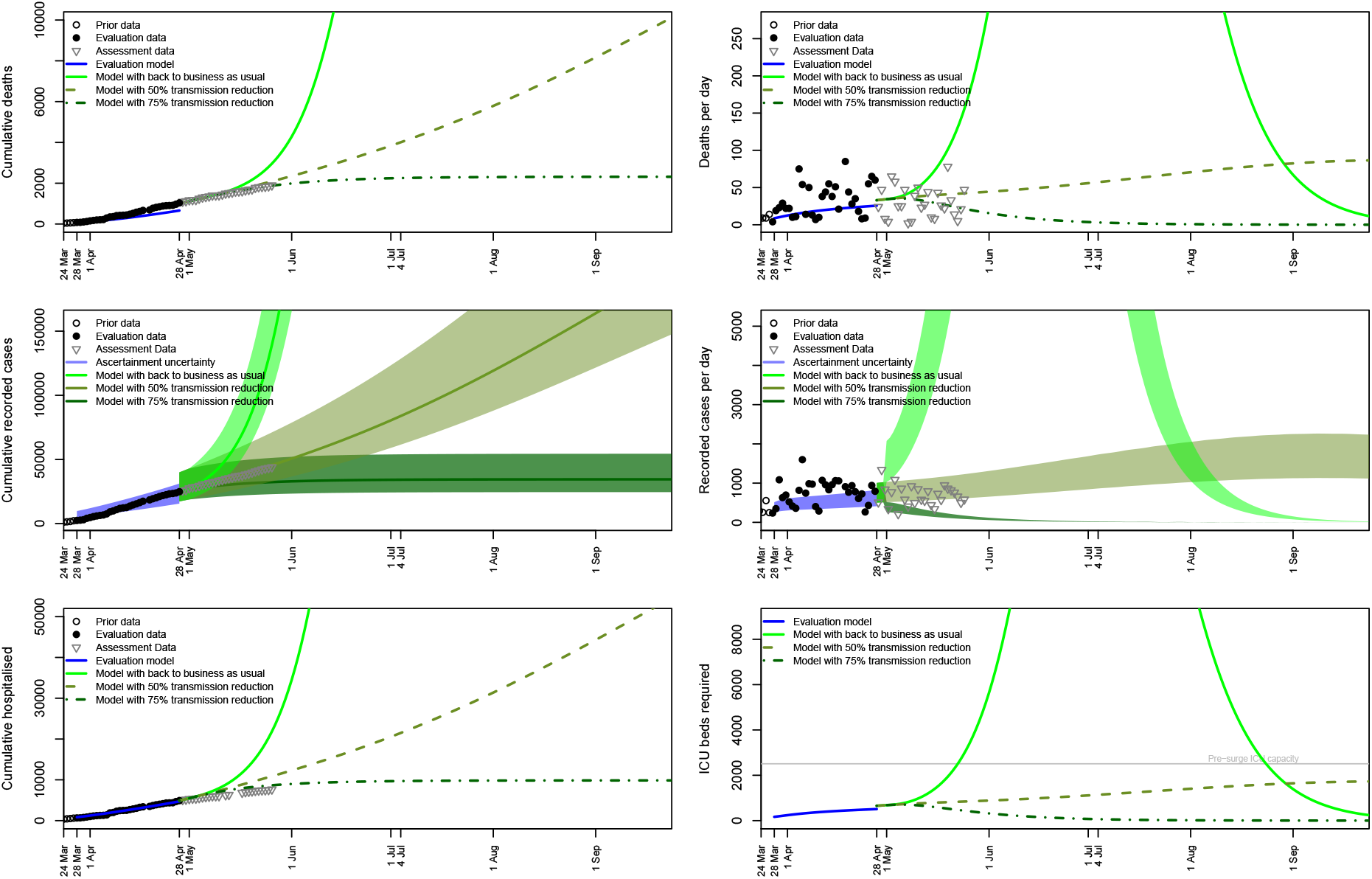
Medium-term COVID-19 projections for the state of Georgia from March 28, 2020 to September 24, 2020. The evaluation model (with 50% social distancing) was initialized on March 28 and recalibrated on April 28 for projections to September 24. Forecasting scenarios represent a return to business as usual, continued intervention, or increased intervention. Case numbers are down-projected by a factor of 5–10*×* to compare with recorded data from testing. a) Cumulative deaths b) deaths per day c) cumulative cases recorded d) cases recorded per day e) cumulative hospitalizations f) number of ICU beds required. Pre-surge ICU bed capacity is highlighted with a horizontal line.

## 4 Discussion

The model framework we developed here incorporates commuting and spatial distributions of population, age-structure and disease-structured epidemiological compartments. Additionally, the model explicitly accounts for asymptomatic carriers and subacute and critical hospitalizations. As such, the MAGE framework is more complex than simple SIR compartmental models (e.g. [7]), and contains explicit information relevant for policy making, but is less complex than other more detailed approaches, e.g., agent based modeling approaches [6]. MAGE is similar to other metapopulation frameworks, like GLEaM [5], albeit in a a deterministic rather than stochastic modeling framework and with a focus on withinlocale rather than between-locale dynamics (e.g., [9]). A potential benefit of the MAGE framework is that it remains reasonably tractable both philosophically – it is possible to see and understand the rules that drive epidemiological dynamics; but also computationally. These characteristics may be appealing for policy makers [32] and for future modeling efforts [33].

We recognize that public awareness of and policies implementing physical distancing – as well as job losses associated with COVID-19 – have and will continue to alter personal mobility patterns. Additional data is required to attempt to constrain how commuting patterns (as assumed here) may be altered by COVID-19 at local scales. For example, counties with extraction, manufacturing, utilities, business management, agriculture, retail trade, transportation and warehousing, and wholesale trade have reduced their mobility the least [34]. Conversely, counties with high levels of finance and insurance, professional, scientific and technical employment have reduced their mobility the most [34]. Exploring epidemiological dynamics in the context of different socio-economic groups and dynamic behavioral change [35, 31] are important future challenges.

As mentioned, forecasts for Georgia with a 50% reduction in transmission rate suggest in excess of 10,000 fatalities by the end of September 2020 (Figure 6, [11]). Currently, there are no proven therapies for COVID-19 [36]. Social distancing has been the primary public health intervention to reduce spread of COVID-19, enhanced and supported by local and national stay-at-home orders. However, this type of intervention strategy may fail if it is relaxed too soon [12, 37, 38] and is already coming into scrutiny by members of the public given the associated socioeconmic and ancillary health costs. Other interventions such as serological shielding [10], digital contract tracing [39] and COVID-19 vaccinations [40] hold promise when they can be integrated as part of strategic decision making.

Our results suggest that the implementation of physical (social) distancing policies have potentially already saved thousands of lives in Georgia alone, and if sustained, are likely to save tens of thousands of lives in the long term. If Georgia remains on the trajectory of a 50% reduction in baseline transmission rates we suggest that the epidemic dynamics will have a shape consistent with a long-term plateau, rather than a sharp epidemic peak. This may be a hallmark of feedback due to heightened social-awareness of the epidemic [11]. However, relaxing physical (social) distancing interventions are likely to lead to an exponential increase in cases, hospitalizations and fatalities. There are opposing tensions between holding down lock-down to reduce disease transmission, and economic, social and personal wellbeing of individuals and businesses [29]. Moving forward Georgia must carefully monitor viral spread as the population remains immunologically naïve at this time. COVID-19 will remain an ongoing threat in Georgia, and globally – potentially into 2021 or beyond [38]. Multiple sustained interventions are likely necessary to protect the general population and safeguard medical resources.

## Data Availability

All simulation and codes used in the development of this manuscript are available at
https://github.com/WeitzGroup/MAGEmodel_covid19_GA/

## Author contributions

SJB and JSW developed the initial concepts and designed simulation scenarios. SJB, MD-M, CA and JSW curated data. SJB designed and implemented the code to perform simulations and visualization. SL and CA provided code materials for mapping data. MD-M performed code review. SJB, MD-M, CA and JSW contributed to analysing and interpreting data. SJB wrote the manuscript. MD-M, SL, CA and JSW edited and revised the manuscript. All authors approved the final version of the manuscript.

## Acknowledgments

We thank K. Carden for sharing ICU data (originally from Kaiser Health news analysis of hospital cost reports filed to the Centers for Medicare and Medicaid services khn.org). We thank those who gave feedback and support to our earlier April 21 and April 28 reports https://weitzgroup.github.io/MAGEmodel_covid19_GA/. Research effort by JSW and co-authors at the Georgia Institute of Technology was enabled by support from grants from the Simons Foundation (SCOPE Award ID 329108), the Army Research Office (W911NF1910384), National Institutes of Health (1R01AI46592-01), and National Science Foundation (1806606 and 1829636). Funding sources had no role or influence on study design, analysis, interpretation, or submission.

## Supporting Information

### S1 State-level model

The system of equations S1 represents a non-spatially resolved epidemic model based on equation set 1 (and previous work [12, 11, 10]). We term this the state-level model:

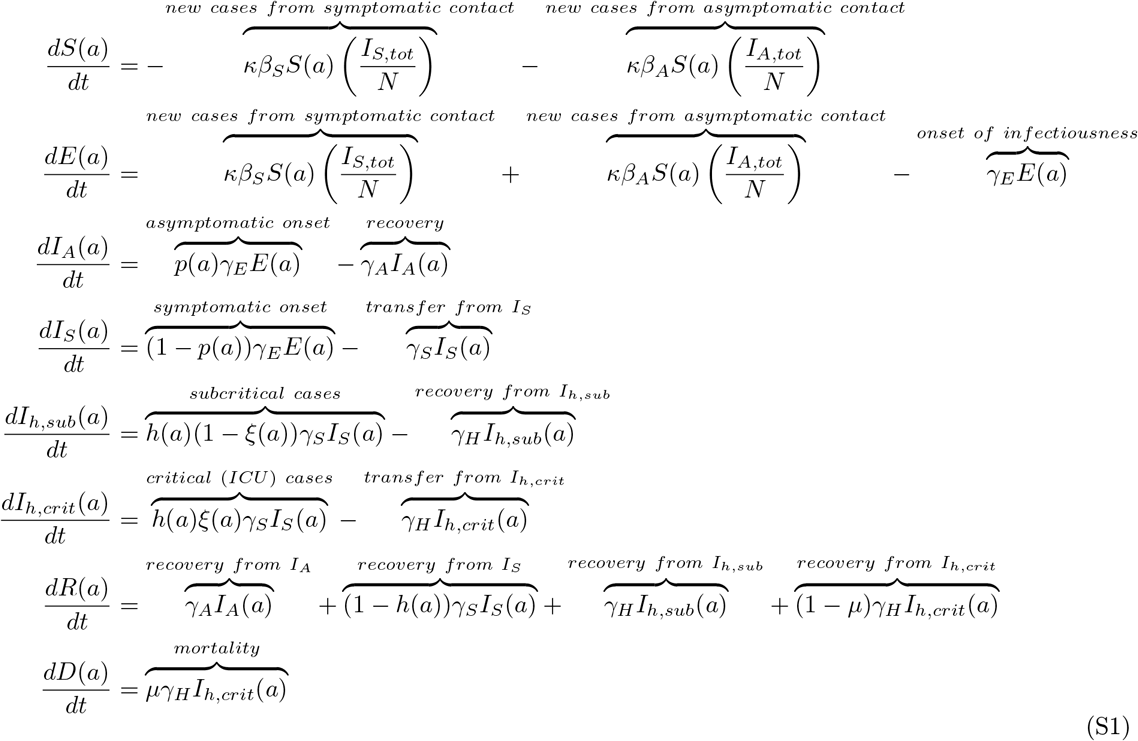

where:

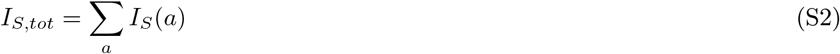

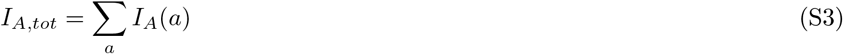

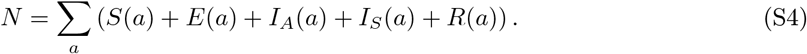

All parameters are the same as in the MAGE model and are given in Tables 1 and 2.

### S2 Construction of the transport matrix

In order to use commuting data in the MAGE model we need to construct what we term the transport matrix, from Origin-Destination datasets. The origin-destination (OD) data shows an absolute number of recorded commutes with origins on the rows and destinations on the columns; and includes data on within-county commutes. We find the total number of commutes originating within each county by summing across the rows of the OD dataset; and use these sums to find the proportions of commutes from each county to other counties. The diagonal elements of this matrix (diag(Mat) denote the proportion of within-county commutes. We replace these diagonal elements with *−*(1 *− diag*(*Mat*)) i.e. the negative of the proportion of out-of-county commutes. Finally, we transpose this matrix (i.e. placing the destinations on the rows and origins on the columns) to create the transport matrix. This structure is made clear in the next section.

### S3 Example of net change in population

Let us define a transport matrix *w* in a network consisting of three nodes (i,j,k):

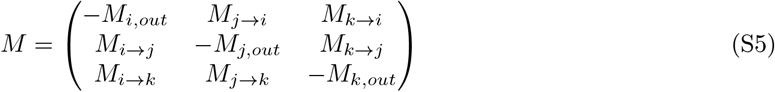

where *M_X→Y_* represents the proportion of the population in node X moving to patch Y. The columns sum to zero as the diagonal element in each column is the negative of the sum of all other column elements. To calculate population net change we multiply the transport matrix by the states of interest – e.g., the susceptible pool in age class 30–39 across the three patches. The calculation for each coupled age-epidemiological state (dropping age notation for convenience) is as follows:

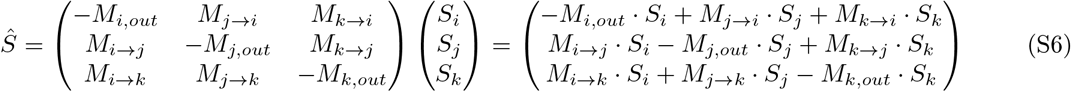

The model diagram (Figure 2) illustrates how commuting is implemented in the context of the epidemiological model. The top panel of Figure 2 shows the core epidemic model and fluxes between epidemic compartments. Note that only five epidemiological groups are assumed to commute between patches: *S, E, I_A_, I_S_, R*. Hospitalized and deceased individuals do not commute. The bottom panel of Figure 2 shows how commuting is represented in a model with three patches i, j and k. For a relative time *w_L_* populations interact in the local phase (in which all patches are assumed to be isolated), representing periods when people are at home or in the local community. However, during the commuting phase which has a relative time of *w_C_*, individuals in the commuting epidemiological groups (*S, E, I_A_, I_S_, R*) are assumed to implicitly move between patches, which can lead to spreading the epidemic. Additionally, this movement is assumed to be age structured – where age categories are derived from the commuting data [19]. Individuals within a patch may stay within the patch or move to other patches (which are not necessarily adjacent) during the commuting phase. Different patches have different commuter patterns, which may also differ by age.

### S4 Evaluating the evaluation phase

Here we explore both the state-level model (SLM) approach described in the main text and an additional method for initializing the outbreak conditions based on county-level simulation (CSM) for the evaluation phase. We find that qualitatively the dynamics of SLM and CSM simulations with the same reduction in social distancing appears similar (Figure S1), though the CSM simulations have a higher baseline than the SLM. We can treat these differences as an analysis of varying the initial conditions. Data suggest that Georgia is on a trajectory most similar to one with a 50% reduction in transmission rates.

**Figure S1:**
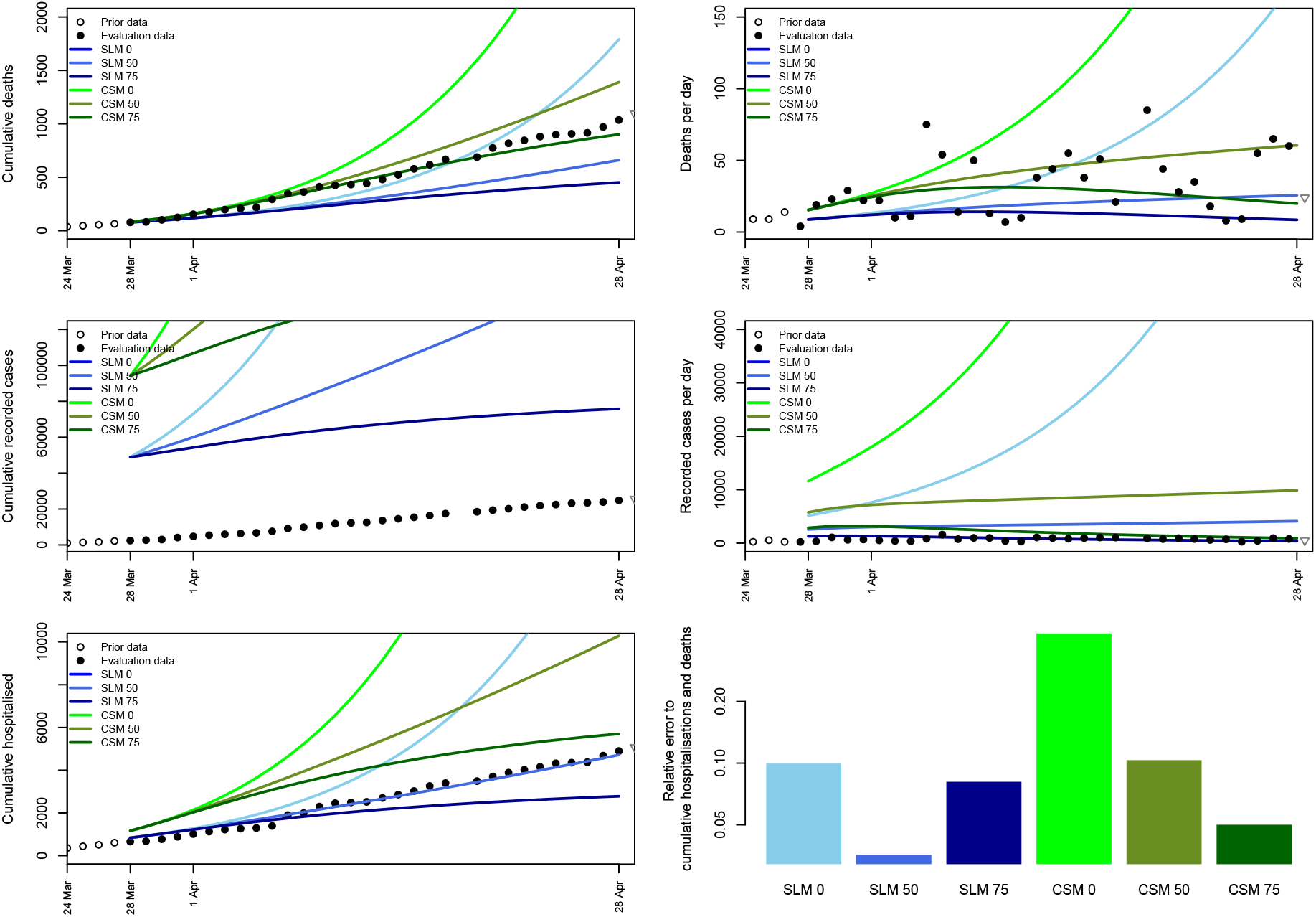
Short-term COVID-19 projections through the evaluation phase for the state of Georgia from March 28, 2020 to April 28, 2020. Projections initialized with the state level model (SLM) are shown in blue, whilst those in green are initialized with the county simulation method (CSM). Lines are shown with no social distancing (0), with a 50% reduction in transmission via social distancing (50) and with a 75% reduction in transmission via social distancing (75). The sum of the relative error as calculated in equation S7 between model simulations and data for cumulative deaths and hospitalizations is shown in the last panel.

#### S4.1 County simulation method

In the county simulation method (CSM), we initialize an outbreak with a single person aged 20–29 in both Fulton County (metro. Atlanta) and Dougherty County (city of Albany). We then simulate the MAGE model forward in time looking county-by-county until the number of reported dead in that county is reached, and record the distribution of people in that county across the different epidemiological states. If no deaths are recorded, we run the simulation until the number of hospitalized cases is reached –assumed to be 20% of the recorded cases; or the fractional equivalent of 0.5 deaths in the county in the event no cases were recorded. When this operation has been performed for each county, we patch together the distributions across epidemiological states county-by-county and use this as the initial condition for the model projections.

#### S4.2 Fitting error

As shown in Figure S1 and explained in the main text our models suggest that there are many more cases in the community than are being reported. Fitting models to case data was not useful given under reporting. To evaluate the fit of each simulation we used a least squares relative error between recorded cumulative deaths and hospitalizations in the model and GA DPH data between March 28 and April 28 as:

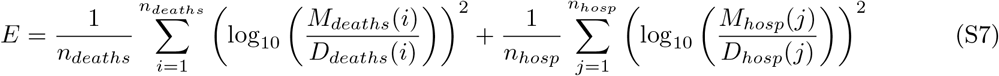

where the number of data points is given by *n_deaths_* and *n_hosp_*, the simulation timeseries are *M_deaths_* and *M_hosp_*; and the data timeseries are *D_deaths_* and *D_hosp_* for cumulative deaths and cumulative hospitalizations respectively.

### S5 County demographics

**Figure S2:**
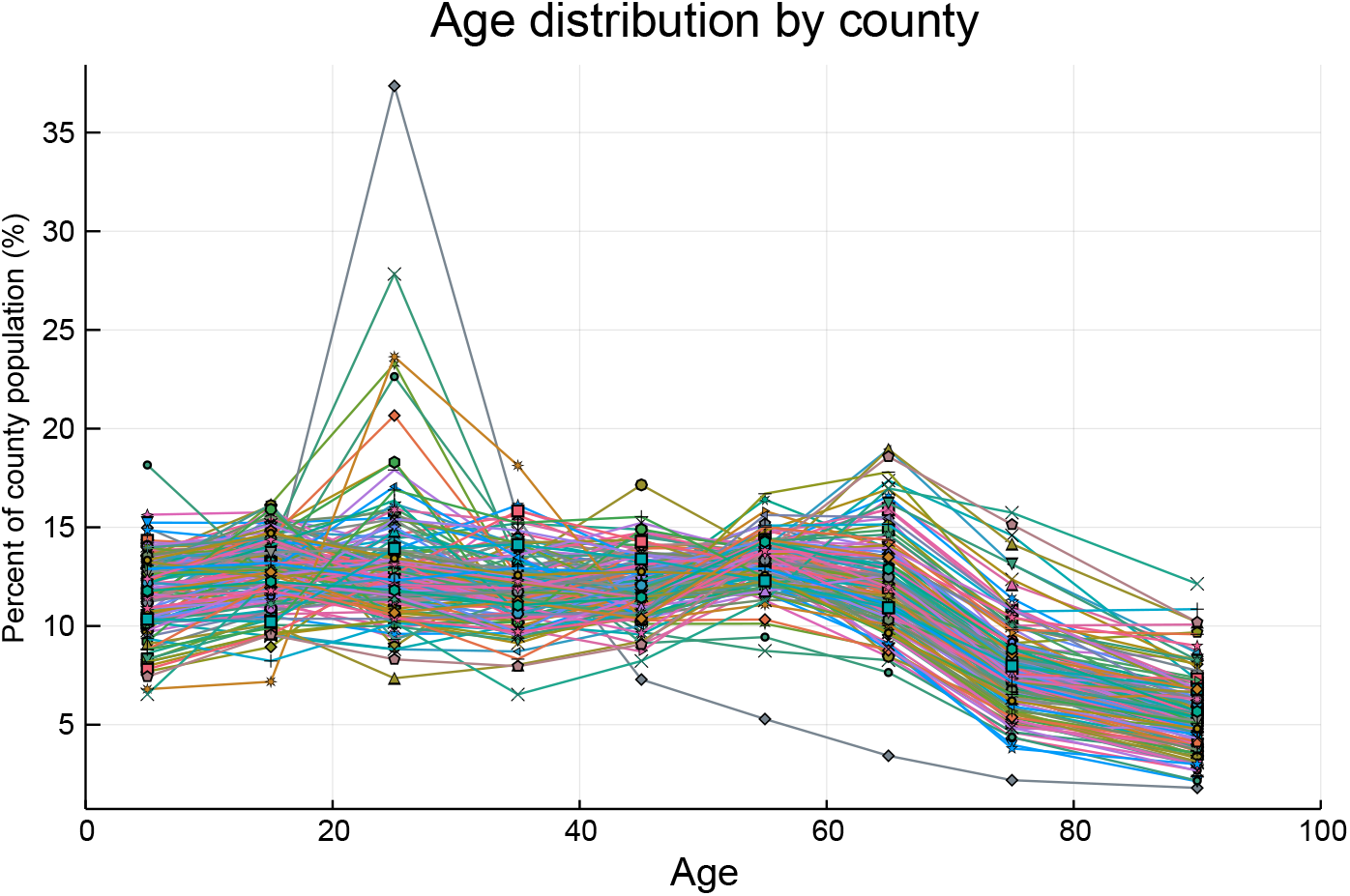
County-level age-structures shown as lines for each of Georgia’s 159 counties per the U.S. Census American Community Survey (2018). Spikes in the 20–29 age class are counties with large universities.

